# Detecting microstructural deviations in individuals with deep diffusion MRI tractometry

**DOI:** 10.1101/2021.02.23.21252011

**Authors:** Maxime Chamberland, Sila Genc, Chantal M.W. Tax, Dmitri Shastin, Kristin Koller, Erika P. Raven, Greg D. Parker, Khalid Hamandi, William P. Gray, Derek K. Jones

## Abstract

Most diffusion MRI (dMRI) studies of disease rely on statistical comparisons between large groups of patients and healthy controls to infer altered tissue state. Such studies often require data from a significant number of patients before robust inferences can be made, and clinical heterogeneity can greatly challenge their discriminative power. Moreover, for clinicians and researchers studying small datasets, rare cases, or individual patients, this approach is clearly inappropriate. There is a clear and unmet need to shift away from the current standard approach of group-wise comparisons to methods with the sensitivity for detection of altered tissue states at the *individual* level. This would ultimately enable the early detection and interpretation of microstructural abnormalities in individual patients, an important step towards personalised-medicine in translational imaging. To this end, Detect was developed to advance dMRI-based Tractometry towards single-subject analysis. By: 1) operating on the manifold of white matter pathways; and 2) learning normative microstructural features to better discriminate patients from controls, our framework captures idiosyncrasies in patterns along brain white matter pathways in the individual. This novel approach paves the way from traditional group-based comparisons to true *personalised radiology*, taking microstructural imaging from the bench to the bedside.

## Introduction

Tremendous progress has been made over the last decade in the non-invasive characterisation of tissue microstructure using diffusion MRI (dMRI). In the brain, for example, information about the structural architecture of the white matter (WM) can be obtained by probing the random motion of water molecules^1^ and acquiring multiple MR images with different diffusion-sensitisation properties. The ability to derive semi-quantitative features such as fractional anisotropy (FA) or mean diffusivity (MD)^2^ or to virtually reconstruct white matter pathways with tractography^3^ has had a huge impact on the ability to distinguish between typical and atypical brain structure *in vivo*, in health and disease^4^. However, prediction modeling (or case-control), in which a *group* of M patients with the same disease is compared with a group of N matched controls, is not well suited to clinically-heterogeneous groups (e.g., autism spectrum disorder, neurological disorders^5^ and rare cases). Despite decades of progress in the research domain, the primary clinical use of dMRI is in emergencies (e.g., diagnosing acute ischaemic stroke or grading and monitoring of tumor invasion). Several studies have shown success in identifying subtle but important microstructural changes at the individual patient-level^6^, showcasing the potential of dMRI to be applied on a broad yet finer scale. Yet, there is a scarcity of dMRI frameworks for single-subject analysis (i.e., 1 patient vs *N* controls). We have therefore reached a hiatus in the characterisation of WM microstructure in disease. There is an urgent yet unmet need for a paradigm shift from group-wise comparisons to individualised diagnosis in computational neuroscience, i.e., detecting whether (and where) the tissue microstructure of a *single* participant is abnormal^7^; not only would this greatly facilitate the study of clinically-heterogeneous groups^8, 9^, it would also facilitate the study of rare diseases and true clinical adoption (i.e., making a diagnosis/prognosis in an individual patient).

In comparing microstructural properties between groups, many computational pipelines adopt a *Tractometry* approach^10, 11^, i.e., mapping measures along pathways reconstructed via tractography, either by averaging along the whole tract^12^, or a segment thereof^13^–16. Along-tract profiling has been applied previously to investigate various brain conditions^10, 17^–19. The main advantage here is that image registration can be avoided as tractometry is performed in each subjects’ native space. There are, however, some limitations. First, most analyses treat tractometry measures from specific pathways as independent measures. This univariate approach has the potential to obscure key relationships *between* different tracts. Focusing on any particular anatomical location therefore increases the risk of losing the full picture. While individual pathways can appear normal in isolation, by considering them as an ensemble (e.g., as part of a network), any such *inter*-tract relationships could collectively help to identify outliers. Second, when analyzing multiple measures (even when derived within the same tract), statistical analysis is hampered by: (i) the multiple comparisons problem; and (ii) any covariance between measurements^12, 20^. Here multidimensional approaches can increase statistical power by combining the sensitivity profiles of independent modalities^20–22^.

The unsupervised multivariate framework proposed here uses state-of-the-art machine learning to approach high-dimensional data non-linearly and improve accuracy and precision over traditional anomaly detection. Normative modeling is an emerging statistical framework that aims to capture variability by comparing individuals to a normative population^8^. Current efforts to apply normative modelling in neuroimaging, however, have so far relied on voxel-based methods, which are suboptimal for WM, whereas reconstructed tracts offer a more intuitive manifold. The framework described here moves significantly beyond dMRI group-level analysis techniques, and uses data-driven normative modeling to identify and localise anomalous tract-profiles at the individual level.

For demonstration purposes, a deep autoencoder was trained to learn normative sets of features derived from healthy tract-profiles in 3 independent datasets and 1 reproducibility dataset^23^. To assess generalization, the framework was then applied to single participants with a range of neurological and psychiatric disorders, including: children and adolescents with copy number variants (CNVs) at high risk of neurodevelopmental and psychiatric disorders; patients with drug refractory epilepsy; and patients with schizophrenia (SCHZ). We compared the performance of our new approach with 1) a conventional Z-score distribution approach; and 2) PCA combined with the Mahalanobis distance (a widely used approach in cluster analysis and classification techniques)^16, 19, 22^.

## Results

### White matter anomaly detection in CNV participants

#### Discriminating power

First, we investigate individual differences in WM microstructure in children with copy number variants (CNVs) at high genetic risk of neurodevelopmental and psychiatric disorders^24^, which are relatively *rare* and therefore challenging to recruit for research imaging studies^25^. For all four microstructural metrics (FA, MD, RISH0 and RISH2 - see Methods), the autoencoder approach was better at identifying CNV subjects as outliers, providing substantially higher sensitivity-specificity trade-offs (Fig. 2, left) than the z-score and Mahalanobis-based approach. In particular, the RISH0 feature showed higher discriminating power (AUC: 0.86 ± 0.06) compared with mean univariate z-score (AUC: 0.53 ± 0.06) and multivariate Mahalanobis distance (AUC: 0.61 ± 0.09). In comparing the RISH0 group distributions, anomaly scores derived via the autoencoder were significantly different (KS=0.62, p<0.003, Cohen’s d effect size: 1.39) between the CNV individuals and the typically developing (TD) subjects. In particular, all CNV subjects had an anomaly score larger than the TD mean and 50% of them were larger than the 95^*th*^ percentile of the TD population. In comparison, the difference between the anomaly scores was less pronounce with the PCA (KS=0.38, p=0.2, Cohen’s d = 0.9) and z-score (KS=0.34, p=0.3, Cohen’s d = 0.38) approaches.

**Figure 1.**
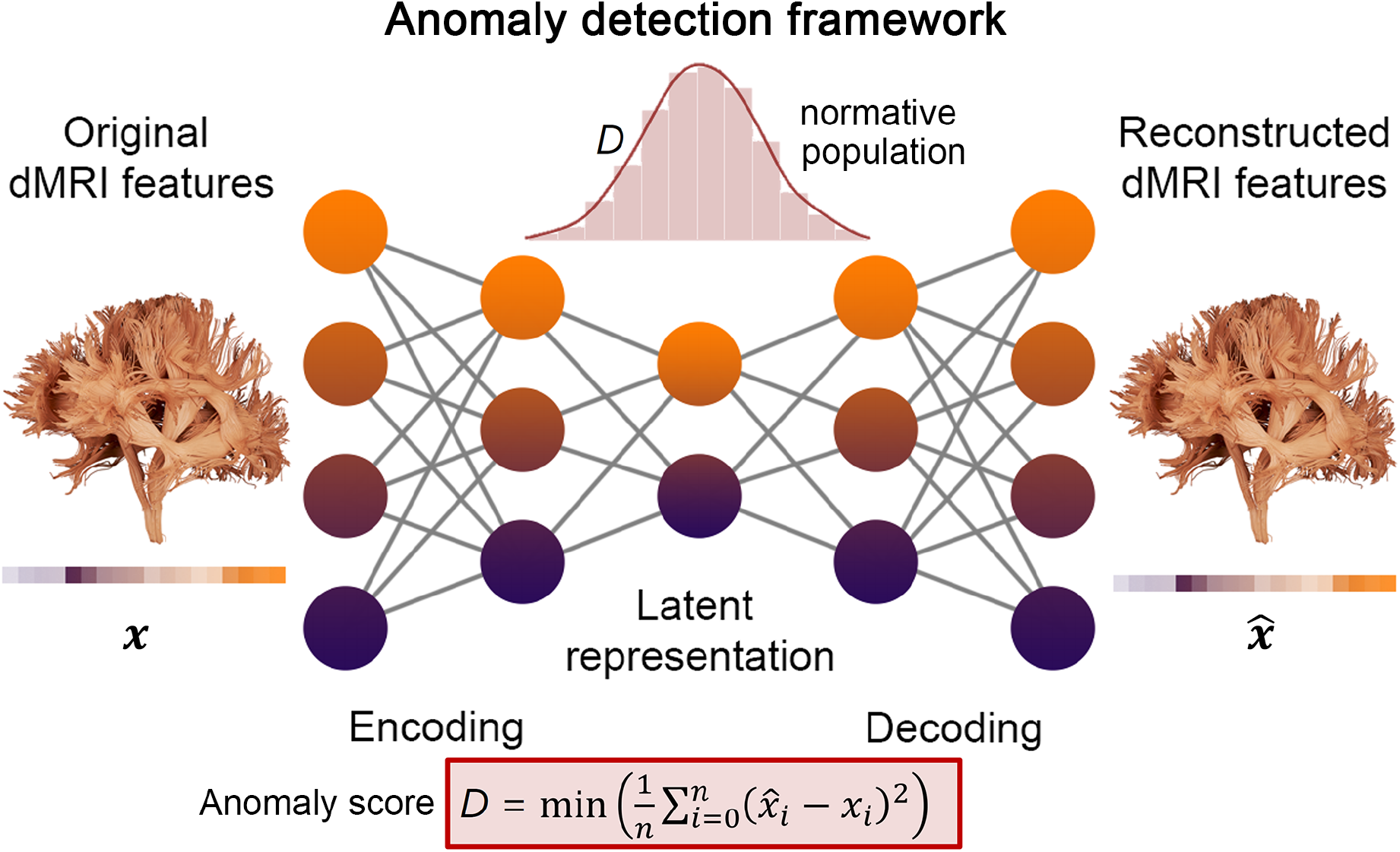
Graphical representation of the proposed anomaly detection framework. The neural network consist in a deep autoencoder symmetrically designed with five fully connected layers. The input and output layers have exactly the same number of nodes as the number of input tracts features (|x|). The goal of the network is to generate an output 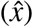 similar to the input (*x*) by minimising the reconstruction error (D). Here, the mean absolute error (MAE) was used as anomaly score. MAE measures the average magnitude of the errors and is derived during testing by computing the absolute differences between the reconstructed micro-structural features 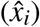 and the raw input features (*x*_*i*_).

**Figure 2.**
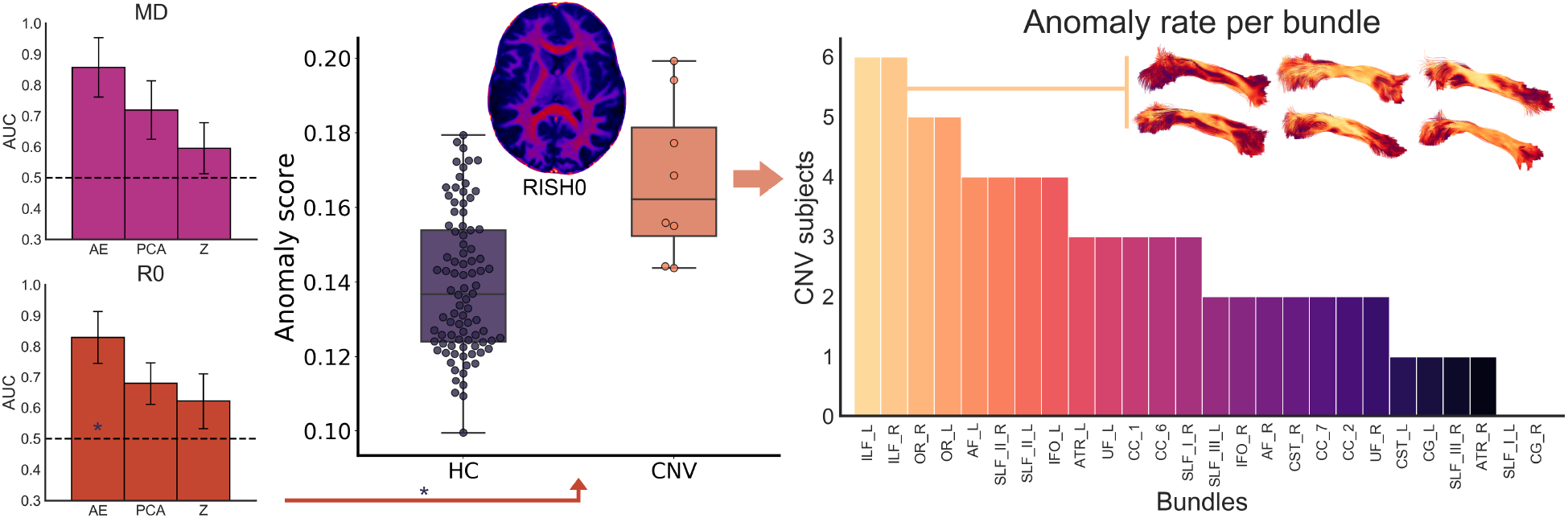
The autoencoder network provides better discriminating power in terms of sensitivity/specificity tradeoffs compared to traditional linear univariate and multivariate approaches (left panel, average AUC over 100 iterations). The RISH0 features show higher reconstruction error for the CNV compared to the HC (precision-recall AUC: 0.45. For comparison, a random classifier would score 0.08). Right: From a group perspective, anomaly rates were mostly observed in the ILF (inset bundles, lateral view), OR and SLF bundles. MD: Mean diffusivity, R0: RISH0.

#### Tract-specific deviations

A key advantage of using deep autoencoders for anomaly detection over traditional PCA-derived approach is its unique ability to relate the anomaly back to the individual elements of the input data. More specifically, the predicted data retains the same dimensionality as the input data and therefore, it is possible to see which feature cannot be accurately recovered by the autoencoder. In contrast, the Mahalanobis distance provides a single summary measure which hinders the interpretation. With deep autoencoders, if a feature has a positive reconstruction error, then one can infer that the network learned a smaller value for that feature than what was provided as input. In the context of the CNV participants, multiple regions were highlighted (by positive reconstruction errors) as deviants from the TD population. Fig 3 reveals a high anomaly rate for various association bundles such as the bilateral inferior longitudinal fasciculus (ILF), optic radiations (OR), and the left superior longitudinal fasciculus (SLF_II). This is in line with current literature where microstructural differences are expected along association pathways, in agreement with psychotic symptoms^26^.

**Figure 3.**
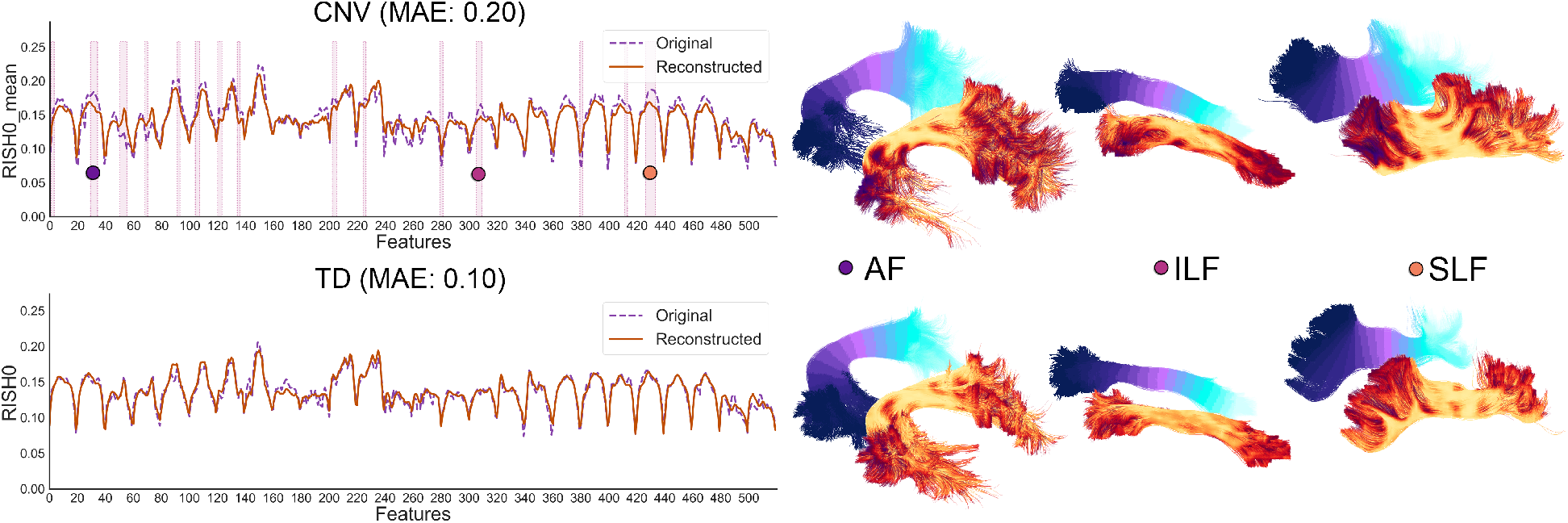
Reconstructed tract profiles of a CNV subject (top) and TD subject (bottom) reveals RISH0 discrepancies along various association bundles such as the arcuate fasciculus (AF), inferior longitudinal fasciculus (ILF) and the superior longitudinal fasciculus (SLF). In particular, the input features for that specific subject were significantly different (red highlighted regions, p < 0.01) than the normal population. MAE: Mean absolute error.

### White matter anomaly detection in epilepsy

Focal cortical dysplasia (FCD), a malformation of cortical development, is the most common etiology in drug-resistant neocortical partial epilepsies^27^. While complete resection is the main predictor of seizure freedom following surgery, a significant proportion of FCDs may be missed with standard clinical imaging protocols^27^ and the seizure generating network may extend far beyond the visible dysplasia. Diffusion MRI contrast enhances the sensitivity of MRI to differences in the brain, but has only been reported at the group level^28^. Here we demonstrate two key advantages of the deep autoencoder approach in a clinical context. First, it succeeded in detecting white matter anomalies that a conventional Z-score based approach has missed, potentially due to hidden interactions between the features; second, detection of abnormal microstructural features away from putative seizure onset zone, as demonstrated in the first example, may contribute to the mapping of epileptogenic networks in individuals. Thus, while the examples shown here had radiological changes detectable with T2-weighted sequences, the method could potentially be extended to cases of “MRI-negative” partial epilepsy increasing the diagnostic yield.

Subject 1 is a young adult female with seizures described as a fuzzy painful sensation in the torso rising up to the head associated with mumbling sounds, occurring 2-5 times per day. Scalp video-EEG showed left temporal inter-ictal epileptiform discharges and left temporal EEG onset. Clinical imaging demonstrated a small area of cortical-white matter junction blurring in the laterobasal left temporal lobe associated with a transmantle area of T2 hyperintensity, suggestive of FCD type II^29^. Neuropsychological assessment was concordant with a left temporal deficit, additionally revealing preserved mesial structures manifesting in relatively preserved verbal memory performance. Subsequent stereo-EEG (SEEG) implantation confirmed ictal onset and prominent interictal discharges from neocortical contacts immediately behind the MRI lesion; in addition, neocortical discharges were seen in SEEG contacts close to the temporal pole. Five tracts of possible relevance were interrogated (Fig. 4). Microstructural anomalies were identified along the left inferior longitudinal fasciculus (ILF) and optic radiation (OR) in the immediate proximity of the T2-weighted changes corresponding to SEEG contacts with maximal ictal EEG changes. Anomalies in the temporal portions of the left inferior fronto-occipital (IFO) and uncinate fasciculi (UF) pointed towards the temporal pole corroborating the SEEG findings that despite normal clinical MRI this area was a part of the seizure network. Based on the clinical findings, the patient proceeded to have resection with histology consistent with FCD.

**Figure 4.**
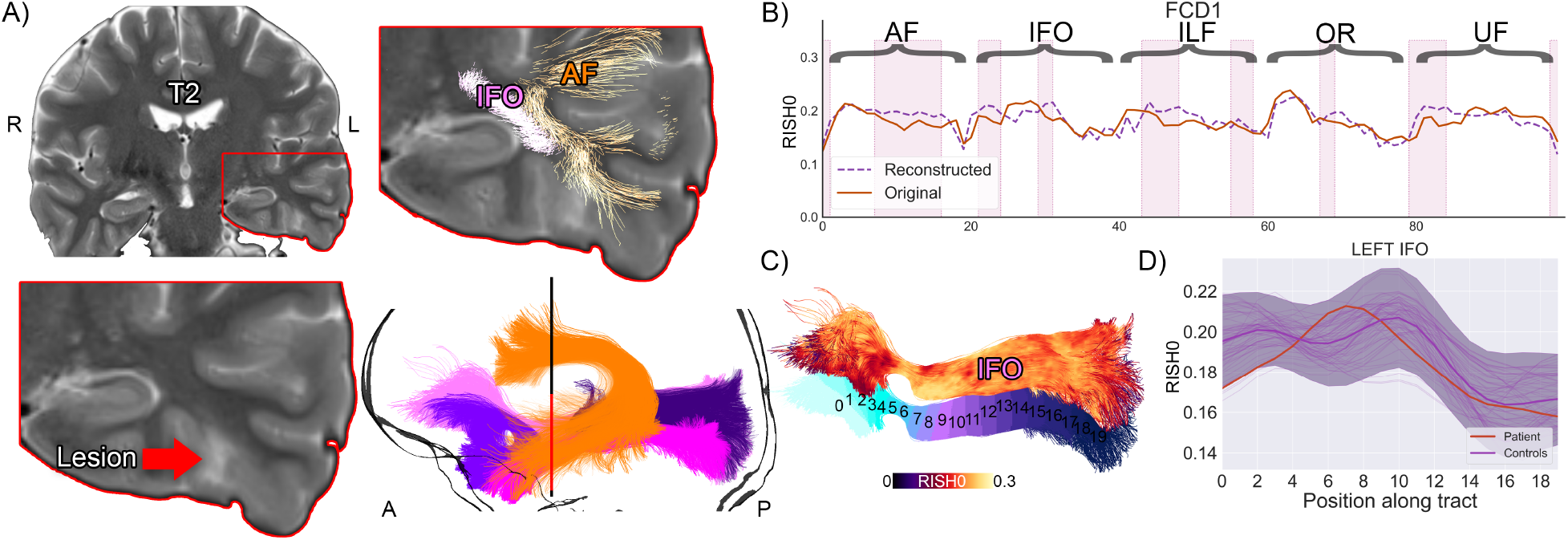
Subject 1 overview. A) T2 hyperintense lesion located at the base of the skull in the temporal lobe. Several pathways with anomalies interdigitate in the vicinity of the lesion. Although the IFO signal did not extend beyond the shaded area (C, D, +/-1 z-score), the proposed anomaly detection framework identified abnormalities in that region (B, pink areas, bold orange line: original tract-profiles, dotted purple line: reconstructed representation learnt from the network).

Subject 2 is an adult female with focal onset seizures since childhood occurring daily with episodes of loss of contact, grimacing and limb stiffening hypermotor movements, including clutching at nearby object on the left side. Scalp video-EEG findings were consistent with frontal onset seizure semiology. Clinical MRI showed blurring of the cortical-white matter junction between the right posterior superior frontal gyrus and the adjacent precentral gyrus, and a transmantle sign on T2/FLAIR from the cortex reaching all the way to the lateral ventricle, consistent with FCD type II. Subsequent stereo-EEG recordings demonstrated spatial overlap between primary motor areas and early ictal onset, and hence the patient did not proceed to surgery. Five tracts of possible relevance were interrogated with our framework (Fig. 5). Anomalies were detected corresponding to radiological and electrophysiological findings along the right corticospinal-tract (CST), primary motor (CC4), and superior longitudinal fasciculus (SLF-I) beyond the visible lesion. No anomalies were found along the right cingulum (Cg) and primary sensorimotor (CC5) regions.

**Figure 5.**
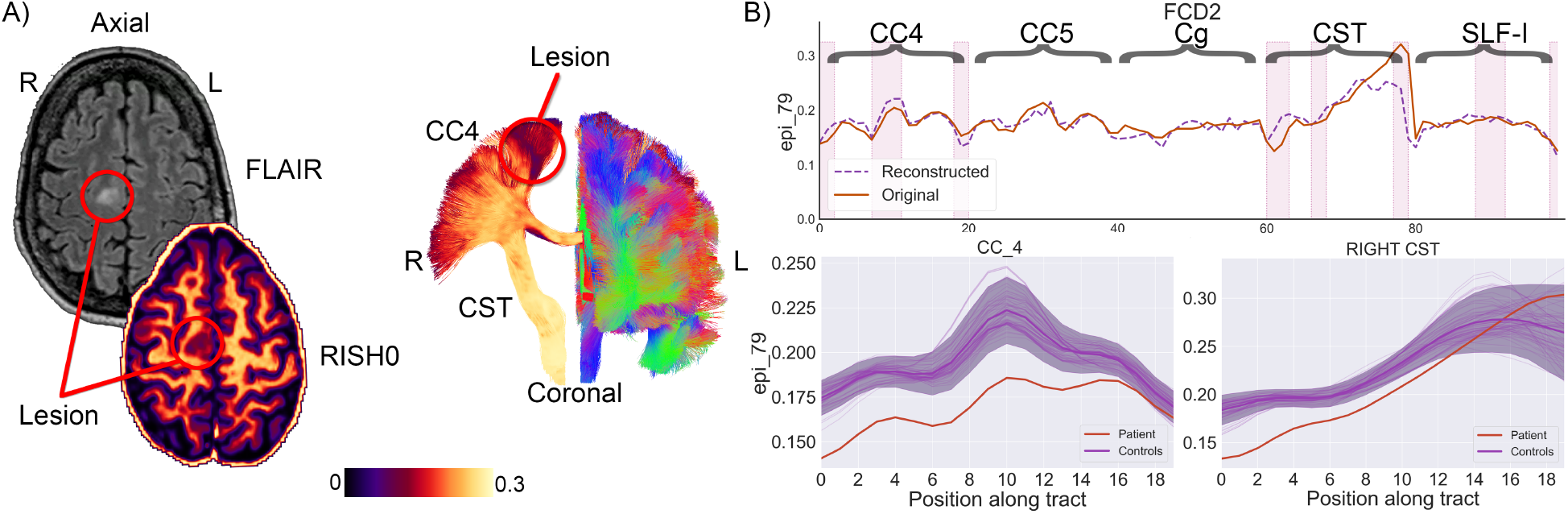
S2 overview. A) The lesion is located anterior to the right primary motor cortex in the supplementary motor area (hyperintense signal on the FLAIR image, hypointense on the RISH map). Tractography show tracts traversing the area (CC4). B) Anomalies were identified in the right CC4, CST, and SLF-Ibundles (top right, pink areas). The bold orange line represents the original tract-profiles whereas the dotted purple line represents the reconstructedrepresentation learned from the network. The z-score approach shows less focused anomaly patterns along the tracts (shaded area: +/-1 Z).

The results are promising, with the tool identifying anomalies in concordance with clinical hypothesis in a single-subject analysis paradigm, testifying to its utility for clinical evaluation. Its additional value is highlighted by its sensitivity to outlying tract segments not detected with the conventional Z-score approach. The *N*=1 approach to detect deep white matter anomalies illustrated here will facilitate the identification of individualised therapy most appropriate to that patient, suggesting additional targets for diagnostic evaluation and possible surgical treatment.

### Linking brain heterogeneity with epidemiological findings in schizophrenia

The extent to which individual clinical variability in schizophrenia relates to microstructural variability remains a key challenge in neuropsychiatry^30^, with most findings being at the group level. Here, the autoencoder approach was better at identifying single SCHZ subjects as outliers (AUC: 0.64 ± 0.06) when compared with PCA (AUC: 0.47 ± 0.08) or z-score (AUC: 0.39 ± 0.06). In comparing these group distributions, anomaly scores derived from the autoencoder were found to be significantly different (t = −2.48, p=0.01, Cohen’s d = 0.47) between the SCHZ individuals and the healthy controls (HC). In particular, 31 of the 43 SCHZ subjects had a anomaly score larger than the HC mean and 9 of them were larger than the 95^*th*^ percentile of the HC population. In comparison, the difference between the anomaly scores was less pronounced with the PCA (t = 1.01, p=0.3, Cohen’s d = 0.18) and z-score (t = 2.05, p=0.04, Cohen’s d = 0.33) approaches. Furthermore, the above chance-level detection rates of the proposed deep autoencoder in SCHZ suggest a successful application of the tractometry-based framework in unsupervised anomaly detection. The significance of these results are even more pronounced considering the challenging task at hand: i.e., where even a supervised support vector machine classifier provides similar accuracy (AUC: 0.65 ± 0.13). We also found that anomaly scores derived from the autoencoder showed higher correlation with the Hopkins anxiety index (Hopkins Symptom Checklist), a widely used screening instrument to study mental illness, than the anomaly scores derived from PCA and Z-score (Spearman’s *ρ* 0.38, p=0.01 vs *ρ* 0.16, p=0.31 and *ρ* −0.12, p=0.45 respectively, Fig. 6).

**Figure 6.**
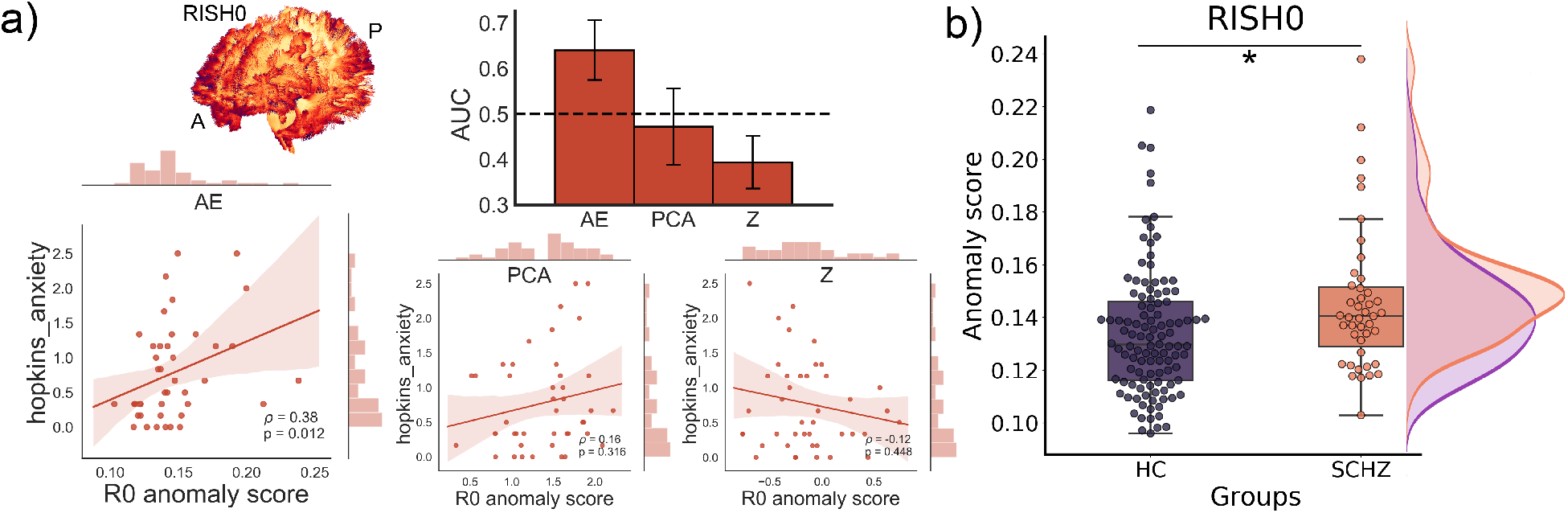
Schizophrenia cohort. a) The autoencoder provides a better discriminating power compared to traditional linear univariate and multivariate approaches with an AUC of 0.64 (bar graph). In addition, the anomaly scores derived from the autoencoder showed higher correlation with the Hopkins anxiety score than the PCA and Z-score correlations. b) The RISH0 features show higher reconstruction error (100 averages) for the SCHZ than the HC (t = −2.48, p < 0.01, Cohen’s d = 0.47).

### Repeatability of anomaly scores and tract-profiles

Using a test-retest dataset (6 subjects, 5 time points), we assessed the repeatability of 1) the input RISH0 tract profiles and 2) the generated anomaly scores by calculating the intra-class correlation coefficient (ICC, two-way mixed, absolute agreement) and the coefficient of variation (CoV). Supplementary Fig. A.1 shows the repeatability of the tract profiles, with the optic radiations (OR) being the most reproducible bundles (mean ICC: 0.95, CoV: 0.03) and the left cingulum being the least reproducible (ICC: 0.66, CoV: 0.06). In terms of anomaly scores, the proposed anomaly detection framework shows reconstruction errors that are reproducible across sessions with an ICC of 0.96 (95% CI: 0.88, 0.99) and a CoV of 0.06.

## Discussion

### Individual characterisation of white matter microstructure and clinical relevance

The framework enabled *subject-* and *tract-specific* characterisation of WM microstructure. By training only on healthy data, our findings revealed that clinical cases (CNVs) were classified as outliers, but not unseen TDs. The framework also outperformed traditional outlier detection mostly due to its ability to handle high-dimensional data non-linearly. This extends the possibility of using anomaly detection in extremely rare cases (as little as n = 1), where group comparisons are otherwise impossible. However, further exploration of input features and hyper-parameters of the model remains to assess the generalizability of the framework and its application to other pathology. We believe that our Tractometry-based anomaly detection framework paves the way to progress from the traditional paradigm of group-based comparison of patients against controls, to a personalised medicine approach, and takes us a step closer in transitioning microstructural MRI from the bench to the beside.

In addition, in an unsupervised novelty detection scenario, the proposed method identifies psychiatric patients from healthy individuals more accurately than mass-univariate normative modeling and a supervised support vector machine classifier. In other words, our approach trained only on healthy participants performs better at detecting abnormal samples than a supervised approach that has full access to the diagnostic labels (as done by^31^). We emphasize that the model has no access to the diagnostic labels during the training phase and thus our anomaly detection approach is fully unsupervised.

In summary, diffusion MRI offers great promise to detect subtle differences in tissue microstructure when applied at the group level. However, the goal of clinical neuroimaging is to be applicable at the individual level. The single case approach proposed here will facilitate the identification of individualised therapy most appropriate to that patient, forming a baseline biomarker for subsequent monitoring through a therapeutic process.

### Generalization of the framework

The key problem in biomarker research is the need for individual prediction/diagnosis. Indeed, advancing knowledge of brain pathology and related cognitive impairment at the individual level is essential for early detection and intervention. Normative models show that, if groups are too heterogeneous, it can be a challenging task to learn characteristics from a given population using supervised approaches, hence the need for unsupervised learning^8^. While the amount of data we can employ in imaging studies is relatively small in comparison with population-based studies, the framework provides a principled method to detect individual differences in tissue microstructure. With the ever-growing amount of dMRI data being acquired, the framework will make less conservative inferences as the number of data points increases. In addition, the framework theoretically can accommodate features beyond and complementary to dMRI. For example, these inputs could include time series from functional MRI (fMRI) and cortical thickness derived from structural T1 imaging. Other applications of the framework include the characterization of microstructural changes in neurological disorders without gross pathology.

### Limitations

Single-subject analysis relies on defining a *normative* brain^8^, which requires significant amounts of healthy-control data. Combining multi-site or multi-scanner dMRI datasets can greatly increase the statistical power of neuroimaging studies^32^ but cross-scanner and cross-protocol variability challenges joint analysis, hence the need for data harmonisation^33, 34^. In addition, tractography still faces significant challenges in the field^35, 36^ and most commonly-available tools can only track reliably within normal-appearing WM. This therefore limits along-tract profiling approaches to core WM bundles only. Providentially, recent ML approaches have shown promise in reproducible tract segmentation across subjects^37^, renewing hope of analysing dMRI data for group studies.

### Novelty

Autoencoders can capture non-linear interactions between the input features and achieve improved feature extraction compared to Principal Component Analysis (PCA) by learning a self-representation of their inputs through a low-dimensional layer (Fig. 1); the goal is to generate an output 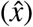 similar to the input (x) by minimising the reconstruction error. This same representation can be exploited for anomaly detection by analysing deviations in the reconstruction. Once trained, the network is then presented with unseen healthy tract profiles (for testing) and subsequently exposed to tract-profiles from individuals diagnosed with neurological/psychiatric brain conditions. Our deep learning approach also provides advantages over other statistical approaches for outlier detection as it was recently shown (using fMRI data^31^) that deep unsupervised approaches improve identification of psychiatric patients compared to mass-univariate normative modeling.

Browser-based applications are becoming increasingly popular amongst the computational neuroscience community due to their ease of use^16^. Here, we developed Detect, an open source tool to promote data exploration through interactive visualizations in the browser. This framework enables the detection of abnormalities in clinically-heterogeneous groups or rare cases and ultimately improve diagnosis of neurological and neuropsychiatric disorders. Our aim was to develop and distribute an open-source framework characterize microstructural white matter changes at the individual level. This enables the detection of abnormalities To the best of our knowledge, other tools like AFQ Browser^16^ compare individuals using a linear approach (z-score) that considers each tract-segment independently and ignores potential complex interactions between the features. Those tract-segment are then statistically tested in an univariate manner, and as such the correction for multiple comparisons - required by the typical high dimensionality of dMRI data - will hamper the discriminating power of the analysis^20^. Recently, PCA was employed to acknowledge the multivariate nature of dMRI data^20–22^, but this approach still relies on linear assumptions thereby ignoring possible complex interactions between the features. We believe strongly that the proposed deep autoencoder approach goes hand-in-hand with existing browser-based dMRI analysis frameworks in encouraging reproducible research and data-driven discoveries. We also encourage future users to experiment and tailor the tool to their needs.

## Methods

### Data acquisition & preprocessing

#### CNV dataset

Diffusion MRI data were acquired from 90 typically developing (TD, age 8-18 years) and 8 children with copy-number variants at high risk of neurodevelopmental and psychiatric disorders (CNVs, 2x 15q13.3 deletion, 2x 16p11.2 deletion, 3x 22q.11.2 deletion and 1x Prader-Willi syndrome) and no apparent WM lesions (age 8-15 years). Data collection procedures for the TD and CNV groups were approved by the Cardiff University School of Psychology and School of Medicine Ethics Committees, respectively. Written informed consent was obtained from all participants and legal guardians. Images were acquired using a Siemens 3T Connectom MRI scanner with 14 b0 images, 30 directions at b = 500, 1200 s/mm^2^, 60 directions at b = 2400, 4000, 6000 s/mm^2^ and 2 × 2 × 2mm^3^ voxels (TE/TR = 59/3000 ms, Δ/*δ* : 24/7 ms). Each dataset was denoised^38^ and corrected for signal drift^39^, motion and distortion^40, 41^, gradient nonlinearities^42^, and Gibbs ringing^43^. Next, rotationally-invariant spherical harmonics (RISH) features^33^ were derived for each subject using the b = 6000 s/mm^2^ shell to maximise sensitivity to the intra-axonal signal^44^ (0^*th*^ and 2^*nd*^ orders only, RISH0 and RISH2 respectively). In addition, diffusion tensors were generated using an in-house non-linear least squares fitting routine using only the b ≤ 1200 s/mm^2^ data, followed by the derivation of fractional anisotropy (FA) and mean diffusivity (MD) maps.

#### Epilepsy dataset

Diffusion MRI data from 2 epilepsy patients with focal cortical dysplasia (FCD) were acquired on a Siemens 3T Connectom MRI scanner with 60 directions at b = 1200, 3000 and 5000 s/mm^2^ and 1.2 × 1.2 × 1.2mm^3^ voxels (TE/TR: 68/5400 ms, Δ/*δ* : 31.1/8.5 ms). In addition, 15 healthy controls (HC) ^1^ (age 21-41 years) from the computational diffusion MRI harmonization database were used^32^. Data collection procedures for the HC and FCD groups were approved by the Cardiff University School of Psychology and School of Medicine Ethics Committees, respectively. Written informed consent was obtained from all subjects. Each dataset were corrected for Gibbs ringing^43^, signal drift^39^, motion and distortion^40, 41^, and gradient non-linearities^42^, Next, RISH features^33^ were derived for each subject using the b = 5000 s/mm^2^ shell.

#### Schizophrenia dataset

Diffusion MRI data from the UCLA Consortium for Neuropsychiatric Phenomics^46^ was downloaded from the OpenNeuro platform openneuro.org/datasets/ds000030/versions/00016, which also contains demographic, behavioral and clinical data. Although more focused on fMRI, the dataset contains dMRI data from 123 healthy controls (HC) and 49 individuals with schizophrenia (SCHZ) amongst other psychiatric disorders. Data were acquired on a Siemens 3T Tim Trio MRI scanner with 1 b0 image, 64 directions at b = 1000 s/mm^2^ and 2 × 2 × 2mm^3^ voxels. Data quality assessment was first performed, resulting in the exclusion of datasets with reduced field-of-view (preventing the reconstruction of white matter bundles in the inferior temporal lobes) and those with significant slice dropout (impacting estimation of diffusion metrics). A total number of 109 HC (age 21-50 years) and 43 SCHZ (age 22-49 years) subjects were used for further analysis. Diffusion data were denoised^38^, corrected for subject motion^40^ and distortion using the anatomical T1-weighted image as reference. Next, RISH features (RISH0, RISH2) were derived for each subject. Finally, diffusion tensors were generated using iteratively weighted least squares in MRtrix^47^ followed by the derivation of FA and MD maps.

#### Repeatability dataset

To assess repeatability, we employed the microstructural image compilation with repeated acquisitions dataset (MICRA,^23^), which comprises 5 repeated sets of microstructural imaging in 6 healthy human participants (3 female, age 24-30 years). Each participant was scanned five times in the span of two weeks on a 3.0T Siemens Connectom system with ultra-strong (300 mT/m) gradients. Multi-shell dMRI data were collected (TE/TR = 59/3000 ms; voxel size = 2 × 2 × 2 mm^3^; bvalues= 0 (14 vols), 200;500 (20 dirs), 1200 (30 dirs), and 2400;4000;6000(60 dirs) s/mm^2^). The same preprocessing as the CNV dataset was applied. Data collection was approved by the Cardiff University School of Psychology Ethic Committee and written informed consent was obtained from all subjects.

### Tractometry

For each dataset, automated white matter tract segmentation was performed using TractSeg^37^ using multi-shell constrained spherical deconvolution (MSMT-CSD,^48^). For each bundle, 2000 streamlines were generated. Tractometry^11^ was performed (sampling FA, MD, RISH0 and RISH2 at 20 locations along the tracts^14, 18, 20^) using a Nextflow architecture provided by SCILPY ^2^. Specifically, individual streamlines were re-ordered for all subjects to ensure consistency using the following order: left-to-right for commissural tracts, anterior-to-posterior for association pathways and top-to-bottom for projection pathways. Next, a core streamline was generated and microstructural metrics at each vertex of the bundle were projected to the closest point along the core. The resulting tract profiles were concatenated to form a feature vector (x).

### Artificial neural network

Our autoencoder implementation consists of a symmetric design of five fully connected layers. The input and output layers have exactly the same number of nodes as the number of input tracts features. The inner layers consecutively apply a compression ratio of 2 by reducing the number of nodes by half, up to the bottleneck hidden layer. Rectified linear units (ReLU) activation was used between the layers to promote sparse activation and *tanh* for the last layer. Using different activation functions in different layers aims at balancing the advantages and disadvantages of the two activation functions. To promote sparsity and reduce overfitting, an activation penalty was imposed to the bottleneck layer using *P*_1_-regularization (10e-5). This is especially best suited for models that explicitly seek an efficient learned representation. 10% of the healthy control data is held out for testing during the training phase (epochs: 25, batch size = 24, learning rate: 1.0e-3, optimiser: Adam, loss: mean squared error). The goal is to generate an output 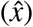 similar to the input (*x*) by minimising the reconstruction error. Here, the mean absolute error (MAE) was used as anomaly score and is defined as:

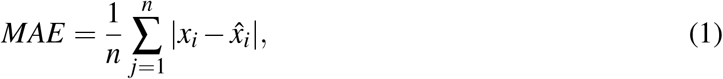

MAE measures the average magnitude of the errors and is derived during testing by computing the absolute differences between the reconstructed micro-structural features 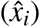 and the raw input features (*x*_*i*_). Due to the heavily-imbalanced group ratio between HCs and patients (i.e., CNV and epilepsy), a bootstrap was implemented to draw random samples of equal sizes from each group. Specifically, a validation set was generated and held-out by combining the patients with a matching random subset of HCs. The rest of the HC data was used to establish a normative distribution. Age regression^49^ and feature normalization (min-max) were performed on the normative training set and subsequently applied to the validation set using a nested approach to prevent information leakage. To derive conservative estimates and assess variations within the model, we repeated this process 100 times and report the mean MAE for each subject. Finally, we compared the sensitivity versus specificity of the anomaly scores using the mean receiver operating characteristic (ROC) area under the curve (AUC) across iterations, and standard deviations used as uncertainties.

### Univariate approach

The z-score (or standard score z = *x* − *µ*)*/σ*) is a way of describing a data point as deviance from distribution, in terms of standard deviations from the mean of the Normal distribution. Here, z-scores were computed for each segment and averaged to derive a subject-specific anomaly score at each iteration (described above). The mean over all iterations was retained as anomaly scores for all subjects.

### Multivariate linear approach

Principal Component Analysis (PCA) was applied to the set of features by restricting the dimensionality to *k* = 3 components. Next, the Mahalanobis Distance (*M*, a multi-dimensional generalization of the z-score that accounts for the relationships between the WM bundles) was used to derive an anomaly score defined as:

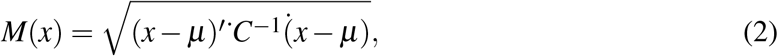

where *x* represents the feature vector of a given subject, *µ* is the vector of mean microstructural metrics for each tract location *s*, and *C*^*−*1^ is the inverse covariance matrix of the input features. The problem of anomaly detection can be seen as a one-class classification problem and therefore, our training data only contains healthy controls to calculate *C*. an *M* score was then derived for all unseen subjects in relation to the healthy control distribution. The same dataset split as aforementioned was used to derive a bootstrapped estimate of *M* for each subject, which was subsequently analysed.

### Support Vector Machine comparison

A supervised support vector machine (SVM) classifier was used for comparisons on the SCHZ dataset. Class weights were set to account for the class imbalance between HCs and patients. The classifier was validated using a repeated (10 times) stratified, 5-fold cross-validation approach in scikit-learn^50^. Optimized parameters were derived for each cross-validation fold using a grid-search approach. Those included the choice of kernel ([radial basis function, linear]), regularization ([1, 10, 100, 1000]) and gamma parameters ([10^*−*3^, 10^*−*2^, 10^*−*1^, 1, 10^1^, 10^2^, 10^3^]).

### Detect interactive interface

Detect is a user-facing tool built upon the Streamlit framework (www.streamlit.io). Users of Detect will input demographic data that consists of comma-separated values (.csv) where each row represents a subject (ID). Example demographics columns include: group, age, gender or clinical scores. The microstructural tractometry data format consist of a. xlsx spreadsheet, where each sheet represents a dMRI metric (e.g., FA, MD, etc.). As per the demographic data, subjects are stacked individually on each rows. The first column denotes the ID of each subject. The remaining columns follow the following convention: *bundle_hemi_section* where *bundle* is the white matter bundle of interest, *hemi* is the hemisphere (i.e., left or right and void for commissural tracts), and *section* is the along-tract portion (e.g., from 1 to 20). The framework offers three main script: *Detect, Inspect* and *Relate*. Detect allows for group comparisons using cross-validated AUCs computed over N iterations, by means of Z-score, PCA or AE (see Supplementary Fig. A.2). The output is a bootstrapped anomaly score for each subject. On the other hand, Inspect allows the user to select a single subject and to compare it with the rest of the population. Here, anomalies in the features are highlighted using a leave-one-out cross-validation approach. In addition, both scripts allow the visualization of tract profiles. Finally, Relate is a simple visual interface to correlate the anomaly scores obtained by the previous commands with clinical scores.

## Data Availability

Detect is an open-source anomaly detection framework for neuroimaging data and will be made available through Github at github.com/chamberm/Detect. The framework is powered by Streamlit www.streamlit.io, an open-source app framework for machine learning and data science. The MICRA repeatability dataset is available at osf.io/z3mkn. TractSeg is available at github.com/MIC-DKFZ/TractSeg. MRtrix is available at www.mrtrix.org. SCILPY is available at github.com/scilus/scilpy. FiberNavigator is available at github.com/chamberm/fibernavigator.

## Data availability

Detect is an open-source anomaly detection framework for neuroimaging data and will be made available through Github at github.com/chamberm/Detect. The framework is powered by Streamlit www.streamlit.io, an open-source app framework for machine learning and data science. The repository will be regularly updated via continuous integration to contain example data (tract-profiles), a Wiki section as well as Jupyter Notebooks with the Python code used to generate the figures in this study. A stable release will also be uploaded to the Python Package Index. The MICRA repeatability dataset is available at osf.io/z3mkn/. TractSeg is available at github.com/MIC-DKFZ/TractSeg. MRtrix is available at www.mrtrix.org. SCILPY is available at github.com/scilus/scilpy. FiberNavigator is available at github.com/chamberm/fibernavigator.

## Acknowledgements

This work was supported by a Wellcome Trust Investigator Award (096646/Z/11/Z), a Wellcome Trust Strategic Award (104943/Z/14/Z), and an EPSRC equipment grant (EP/M029778/1) to DKJ, a Sir Henry Wellcome Fellowship (215944/Z/19/Z) and VENI grant (17331) from the Dutch Research Council (NWO) to CMWT, a Wellcome Trust GW4-CAT Fellowship (220537/Z/20/Z) to DS, and a NIH NICDH fellowship (1F32HD103313-01) to EPR. This study is also supported by the Brain Repair and Intracranial Neurotherapeutics (BRAIN) Unit, funded by Health and Care Research Wales awarded to WPG. The authors thank Prof. Maxime Descoteaux and Jean-Christophe Houde (Sherbrooke Connectivity Imaging Lab) for their useful discussions and code sharing, and thank Adam Cunningham, Joanne Doherty and Marianne van den Bree (Cardiff University) for recruiting the CNV patients.

## Author contributions statement

M.C. developed the framework, conducted the experiments, analysed the results, and wrote the manuscript, M.C., C.M.W.T. and D.K.J. conceptualized the project, S.G. and E.P.R. acquired the paediatric data, M.C. and S.G. pre-processed the pediatric imaging data, G.D.P. and C.M.W.T. developed the pre-processing pipeline, W.P.G., D.S. and K.H procured the epilepsy data, M.C., C.M.W.T. and D.S. acquired and pre-processed the cdMRI and epilepsy data, K.K. acquired the MICRA repeatability data. All authors reviewed and contributed to the manuscript.

## A Supplementary material

**Figure A.1.**
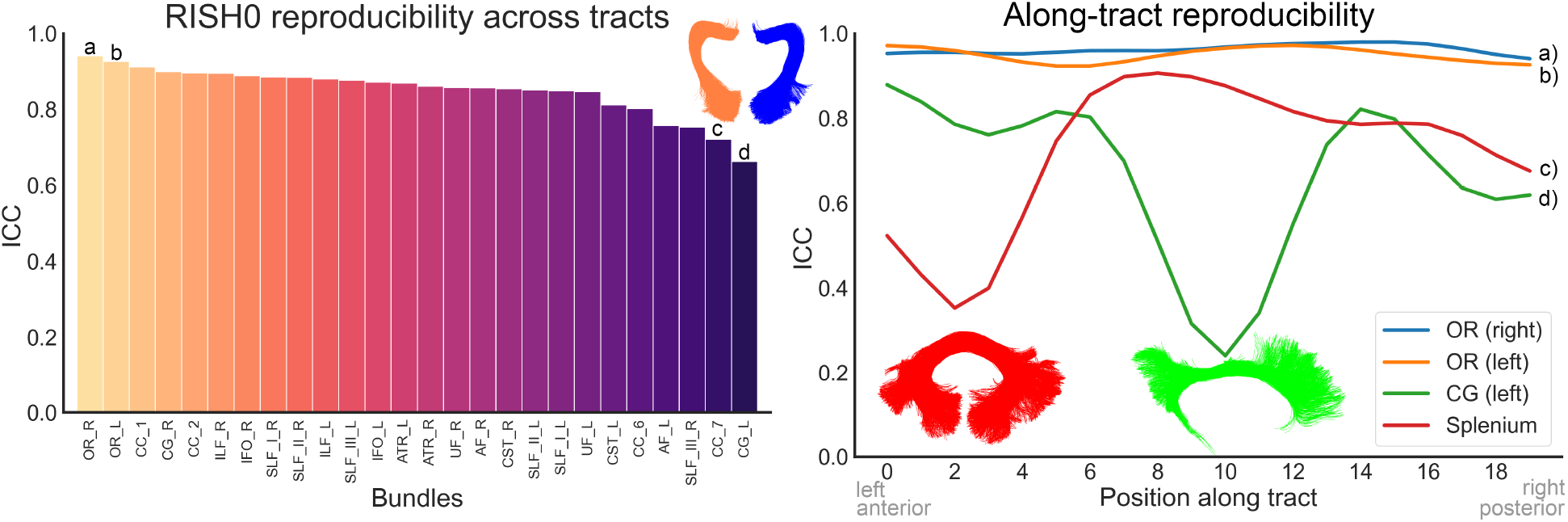
Tract-profile repeatability of RISH0. Repeatability was assessed using the intra-class correlation coefficient (ICC, two-way mixed, absolute agreement) computed over 6 subjects (5 time points). Most of the bundles show excellent repeatability (mean ICC: 0.86, mean CoV: 0.03). In particular, the left and right optic radiations ranked amongst the most reproducible bundles, whereas the splenium and left cingulum show the lowest scores. The along-tract ICC profiles revealed anatomical locations where the ICC was lower in those bundles. a) Optic radiation (right), b) Optic radiation (left), c) Splenium (CC_7), d) Cingulum (left).

**Figure A.2.**
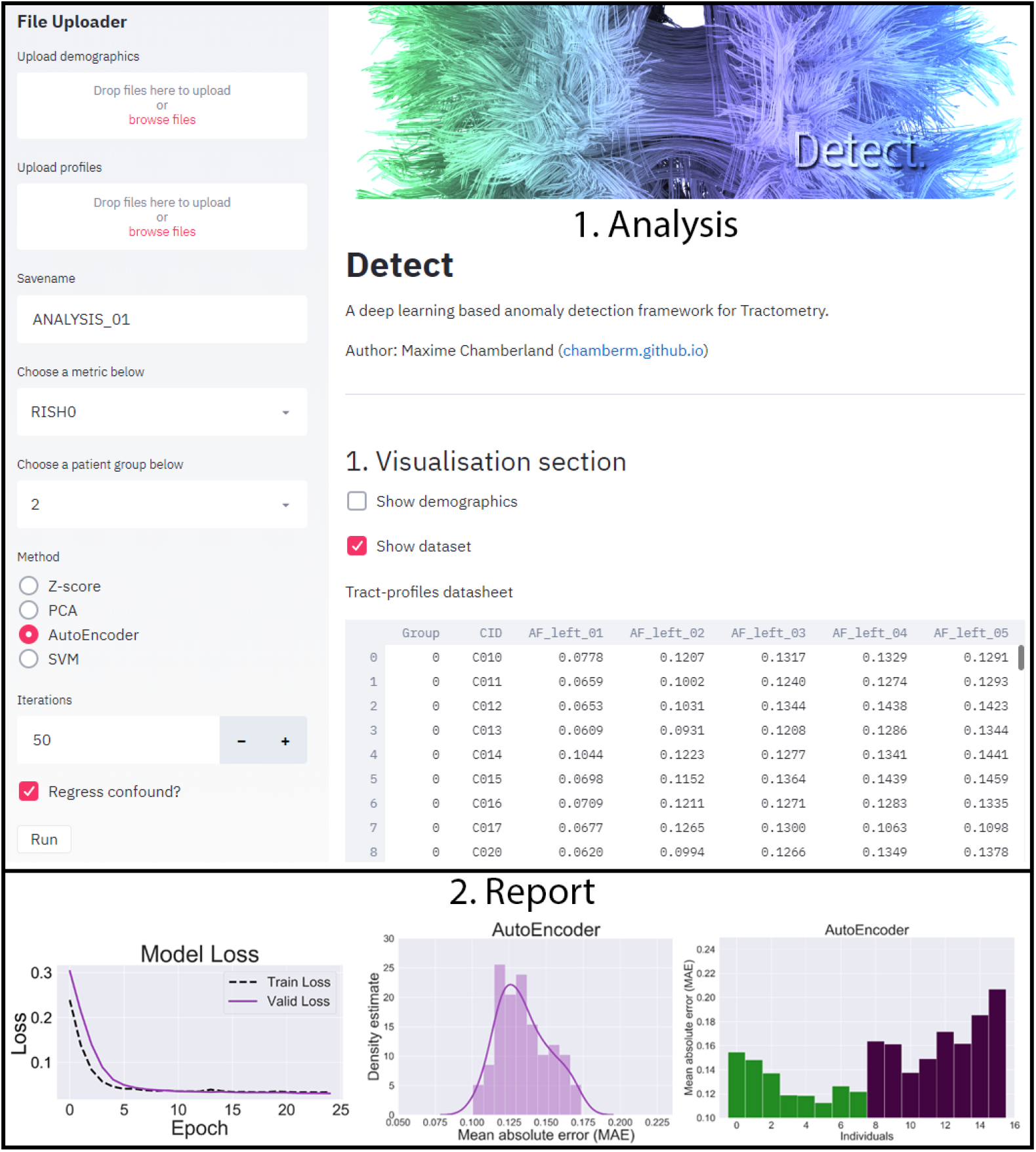
Overview of the proposed interactive application in the browser. The user has access to various setting on the left panel, including the choice of the diffusion metric, the anomaly method, and the number of iterations. The main display updates in real-time during computation (after each iteration), showing the training loss, the distribution of anomaly scores and the ROC results. At the end, a final report with the computed anomaly scores for each subject is saved for further analysis.

## Footnotes

1 A four-fold data augmentation was applied to the HC tract-profiles using a Synthetic Minority Over-sampling TEchnique (SMOTE^45^) resulting in 75 HC.

2 github.com/scilus/scilpy

